# Informative Missingness: What can we learn from patterns in missing laboratory data in the electronic health record?

**DOI:** 10.1101/2022.05.08.22274724

**Authors:** Amelia L.M. Tan, Emily J. Getzen, Meghan R. Hutch, Zachary H. Strasser, Alba Gutiérrez-Sacristán, Trang T. Le, Arianna Dagliati, Michele Morris, David A. Hanauer, Bertrand Moal, Clara-Lea Bonzel, William Yuan, Lorenzo Chiudinelli, Priam Das, Harrison G. Zhang, Bruce J Aronow, Paul Avilllach, Gabriel. A. Brat, Tianxi Cai, Chuan Hong, William G. La Cava, He Hooi Will Loh, Yuan Luo, Shawn N. Murphy, Kee Yuan Hgiam, Gilbert S. Omenn, Lav P. Patel, Malarkodi Jebathilagam Samayamuthu, Emily R. Shriver, Zahra Shakeri Hossein Abad, Byorn W.L. Tan, Shyam Visweswaran, Xuan Wang, Griffin M Weber, Zongqi Xia, Bertrand Verdy, The Consortium for Clinical Characterization of COVID-19 by EHR (4CE) (Collaborative Group/Consortium), Qi Long, Danielle L Mowery, John H. Holmes

**Affiliations:** Harvard Medical School, Cambridge, MA, USA; University of Pennsylvania Perelman School of Medicine, Philadelphia, PA, USA; Northwestern University, Chicago, IL, USA; Massachusetts General Hospital, Boston, MA, USA; University of Pavia, Pavia, Italy; University of Pittsburgh, Pittsburgh, PA, USA; University of Michigan, Ann Arbor, MI, USA; Bordeaux University Hospital, Talence, France; ASST Papa Giovanni XXIII, Bergamo, Italy; Cincinnati Children’s Hospital Medical Center, University of Cincinnati, Cincinnati, OH, USA; Duke University, Durham, NC, USA; National University Health Systems, Singapore; University Of Kansas Medical Center; University of Pennsylvania Health System, Philadelphia, PA, USA; Boston Children’s Hospital, Boston, MA, USA

## Abstract

**Background:** In electronic health records, patterns of missing laboratory test results could capture patients’ course of disease as well as reflect clinician’s concerns or worries for possible conditions. These patterns are often understudied and overlooked. This study aims to characterize the patterns of missingness among laboratory data collected across 15 healthcare system sites in three countries for COVID-19 inpatients.

**Methods:** We collected and analyzed demographic, diagnosis, and laboratory data for 69,939 patients with positive COVID-19 PCR tests across three countries from 1 January 2020 through 30 September 2021. We analyzed missing laboratory measurements across sites, missingness stratification by demographic variables, temporal trends of missingness, correlations between labs based on missingness indicators over time, and clustering of groups of labs based on their missingness/ordering pattern.

**Results:** With these analyses, we identified mapping issues faced in seven out of 15 sites. We also identified nuances in data collection and variable definition for the various sites. Temporal trend analyses may support the use of laboratory test result missingness patterns in identifying severe COVID-19 patients. Lastly, using missingness patterns, we determined relationships between various labs that reflect clinical behaviors.

**Conclusion:** This work elucidates how missing data patterns in EHRs can be leveraged to identify quality control issues and relationships between laboratory measurements. Missing data patterns will allow sites to attain better quality data for subsequent analyses and help researchers identify which sites are better poised to study particular questions. Our results could also provide insight into some of the biological relationships between labs in EHR data for COVID-19 patients.

## Introduction

The increasing availability of electronic health record (EHR) data has led to the burgeoning use of these data in various domains, including the identification of disease phenotypes and the clinical course of disease. Most recently, the EHR has been used as a rich source of data for characterizing the trajectory of the Coronavirus Disease (COVID-19, or simply COVID) that is caused by the SARS-CoV-2 virus. However, it is commonly acknowledged that EHR data often require rigorous processing and cleaning before they are of usable quality, thereby presenting considerable challenges to those using these data for research, quality improvement, or disease surveillance. Issues such as data availability [1–3], data recording or format inconsistencies [4,5], temporal changes in data policies [5,6], poorly standardized free-text [1,6], lack of interoperability between EHR systems [6,7] and diagnostic coding errors [1] all impair the usability of EHR data. Moreover, the most frequently reported barrier to EHR usability is missing data or data which are expected to be in the record but are not [3,4,6,8–12].

There is an important difference in the way that clinicians and biomedical informaticians tend to view missing data. To a clinician, data is considered missing if a laboratory test was supposed to be conducted and its value recorded, but for some known or unknown reason, it is absent from the medical record. Therefore, if a test is measured and recorded once per week according to protocol, and this is carried out without issues, there is no missing data for this particular test. On the other hand, if an informatician is carrying out a time series analysis that requires a measured value each day, that laboratory test will be considered missing for the 6 days it was not measured. This study was originally motivated by the desire to carry out a time series analysis; but with laboratory tests collected at different frequencies, the issue of missing data and how to deal with it needed to be addressed. As we delved into this, we realized that missing data itself could be leveraged to learn more about EHRs, nuances across sites, ordering patterns, and relationships between the labs themselves.

Data is missing in the EHR for two principal reasons. First, a laboratory test might have been ordered, but the result is missing from the record. Although important for ascertaining the quality of reporting systems, characterizing this type of missing data is difficult without access to clinical notes and ordering systems in the EHR. The second reason missing data is that a laboratory test was never ordered, or where a test was ordered and resulted, but for some reason was not resulted for some time or ever again during a hospital stay, and thus a result would not be expected in the EHR. Such missing data should not be considered a direct measure of EHR data quality, since there are many factors, often clinical, that determine when and if a test result is absent from the record. We focus here on this type of missing data, and we propose that the absence of data can be informative, and that patterns of these missing data can be considered as *informative missingness*.

### Patterns of Data Missingness

Missing data is typically characterized according to three commonly accepted *missingness patterns*. The first is *missing completely at random* (MCAR). In this pattern, the missingness of a variable is not associated with any observed or unobserved variables, including the variable itself. An example would be where responses to a survey question about smoking status is not present on some proportion of respondents because the question was asked (or not) in a truly random fashion; in other words there is no nonrandom pattern of missingness. In the second pattern, *missing at random* (MAR), the missingness of a variable is associated with the value of another observed variable. For example, responses to a question on smoking status are dependent on one’s occupation, resulting in a missing value for smoking when a respondent notes that their occupation is in health care. Finally, when data are missing in the EHR, it is often *missing not at random* (MNAR). A variable may be missing because of the value of the variable itself. For example, a smoker may be less likely to answer a survey question about smoking status because they are a heavy smoker. Note that it is not known whether or not the respondent is, in fact, a smoker, and that would not be known because they did not answer the question. In other words, the probability of determining if a respondent is a smoker is depending on the value of the smoking question. For this reason, such missing data are nonignorable, which implies that such data violate assumptions for imputation and need to be considered (encoded) explicitly as missing data for purposes of imputation.

However, we contend that there is a special case of MNAR, where the missing data are *informative*, in this case indicating a clinician’s assumption and a decision not to order a test subsequent to the previous one. This type of missingness has been referred to as structurally missing data [13], in that there is a logical, non-random reason the data are missing. However, we refer to this pattern as *informative-missing not at random* (I-MNAR). In this pattern, the missingness of a variable is dependent on the value of the variable, like MNAR, but may also be influenced by the value of other variables as well, whether they are observed *or* unobserved. It is a pattern commonly seen in the EHR, where once a normal laboratory result is obtained, no further assays of the same type are present in the record. The absent results indicate that the decision not to order the test after the normal result was likely due to the normal value itself, but it could be that the values of other variables (such as other laboratory tests or clinical assessments) are taken into account during the decision-making process. Thus, the absence of laboratory results after a given result is informative, perhaps about the severity of the disease, the availability of the test, practice guidelines, or clinician preferences. An example of this is in [14], where the recording of rheumatoid factor test results in the EHR was found to be missing when a test result was negative. In other words, a test was not ordered because it was assumed that the test would be negative based on a prior result of the test.

### Informative Missing-Not-At Random in EHR Studies

The I-MNAR pattern has been investigated in the literature, albeit under a different nomenclature, typically referred to simply as “informative missingness”. For example, in [15] it was noted that missing data are often correlated with a target variable, such as outcome. Informative missing data has been identified in genotype analysis and genetic association studies [16–21], longitudinal cohort studies [22,23], meta-analyses [24–29], exposure assessment in case-control studies [30], and particularly in studies using EHR data [31].

The goal of this study was to identify patterns of missing laboratory tests that might suggest levels of disease severity or other factors, such as patient sex or hospital characteristics that could influence the availability of laboratory data in the EHR. From this study, we hope to determine if these missing data fit an I-MNAR pattern and interpret this informativeness. Accordingly, we focus here specifically on the laboratory data patterns found in our examination of EHR data in a large international federated data consortium.

### Setting

For this study, we used the resources of the Consortium for the Clinical Characterization of COVID-19 by the EHR (4CE). The 4CE consortium includes 342 hospitals in eight countries with patients who have been hospitalized for COVID. The 4CE uses a federated data model and predictive analytics framework in a hub-and-spoke configuration. Specifically, all 4CE-contributing academic medical centers (spokes) query and standardize EHR data elements using a COVID ontology, and apply analysis locally to their COVID datasets, and then provide aggregate statistics to a coordinating academic medical center (hub). This agile, rapid, and privacy-preserving data sharing approach has efficiently and effectively supported several COVID studies over the last two years [32–37]. We examined EHR data from 15 4CE-participating sites representing 232 hospitals in the United States and Europe comprising 69,939 patients for this study. For sites with multiple hospitals, we assume that practice patterns are similar across the hospitals within those sites.

This manuscript is structured into two main components; First we use patterns in missingness for identification of QC issues; second, we delve deeper into the relationships between patterns of missingness between laboratory measures. After this introduction, we describe our methodology for defining the sample population, the variable set, and the analytic methods used to quantify and describe missing values. Next, we present the results as characterization of the distribution of missing data by laboratory test, stratified by sex, hospital site, and disease severity, across different time periods from admission through 60 days thereafter. We also present the results of an analysis that seeks to investigate the patterns of missingness with regard to pairs of laboratory tests. Finally, we consider a topic model analysis that clusters groups of labs together based on missingness patterns.

## Methods

This retrospective observational study of EHR was reviewed and approved by the ethics and institutional review boards for all participating 4CE contributing sites.

We analyzed laboratory test ordering patterns in EHR data for 69,939 patients from 232 hospitals across three countries from the 4CE consortium for the period of 1 January 2020 through 30 September 2021. 4CE consortium contributing sites are described in **Table 1**.

**Table 1.**
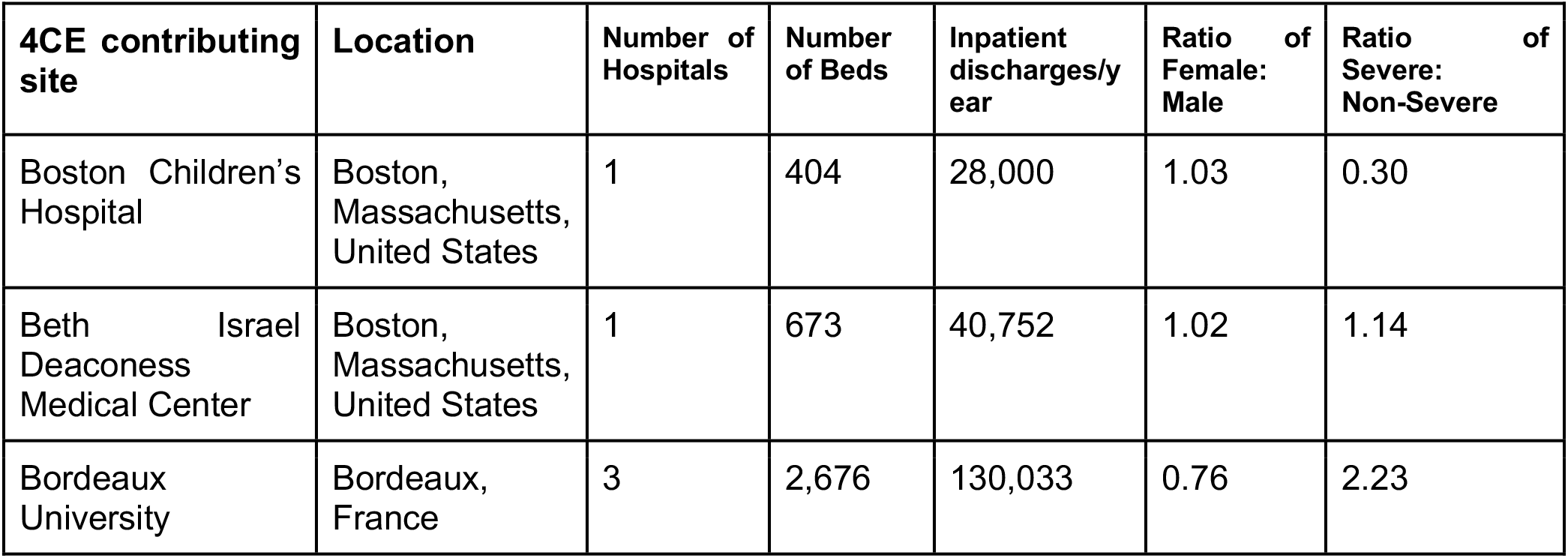
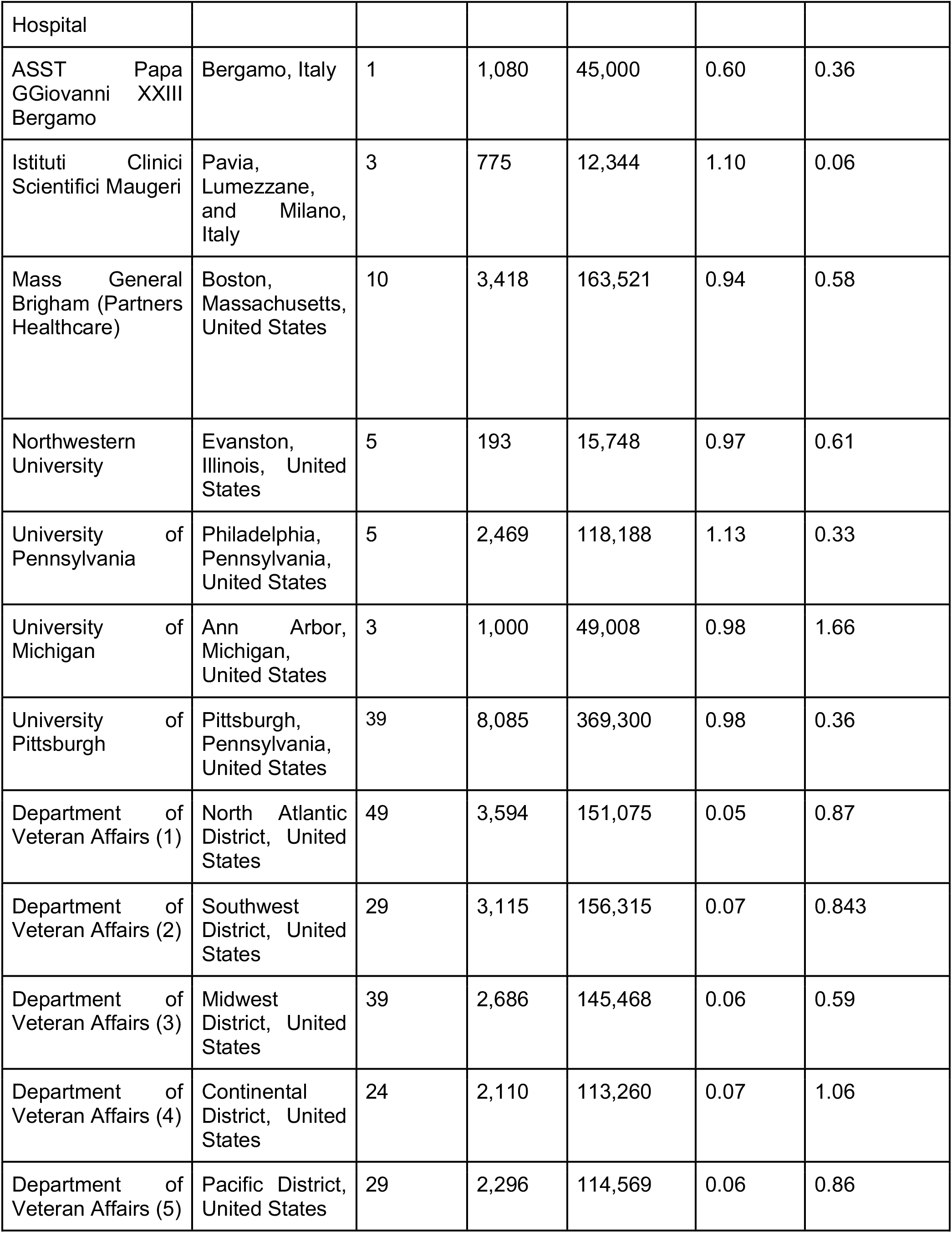
4CE contributing sites.

The inclusion criteria include a positive COVID-19 polymerase chain reaction (PCR) test on or during admission to the inpatient setting. Only data from a patient’s first COVID-19 admission was considered; subsequent admissions were not included in the analysis. We collected results from 16 laboratory tests conducted over the entire admission. We selected these labs because (1) their abnormal values have been associated with worse outcomes among COVID patients in the literature and in our own 4CE mortality risk prediction models and (2) their ability to reflect acute pathophysiology of COVID-19 patients. The clinical significance of these tests and and common ordering practice is described in **Supplementary Table 1** [38–40].

### Quantifying missingness: definitions

A missing laboratory test is logged when no results for the test are available in the EHR for a patient within a time point, usually within a day. Thus, we assume that a missing test result serves as a proxy for a test that wasn’t ordered. The number of missing results per patient will hence be the total number of days without a given laboratory test result for the period of hospital admission. Note that this is oftentimes not how missing data are defined in a clinical setting. Even though it could be routine practice not to order a given laboratory test each day, we still want to capture this information on a per-day basis to understand the ordering patterns of the various labs.

We first investigate the overall number of missing results and the proportion of missing values for each site and lab. The proportion missing for a given patient is defined as the total number of hospital days with no results for a given laboratory test divided by the total number of days admitted. The purpose of this analysis is to elucidate potential quality control issues that might exist in our data; for example, it is standard practice for lymphocytes and neutrophils to be ordered together. Thus, if a site has differing amounts of missing data for these two labs, that may indicate a mapping issue. Other combinations that share this relationship are AST and ALT.

### Quantifying missingness across three selected indicators

We characterized the degree of missingness across variables of *severity, sex*, and *time*. We defined COVID severity by applying an EHR-based algorithm that defines severe patients based on the blood gas results (partial pressure of carbon dioxide or partial pressure of oxygen), medications (sedatives/anesthetics or treatment for shock), diagnoses (acute respiratory distress syndrome or ventilator-associated pneumonia), and procedures (endotracheal tube insertion or invasive mechanical ventilation) [33]. A patient with one or more of the aforementioned data elements was noted as severe; otherwise, the patient was assigned as non-severe. We assess for differences in quantiles of missingness between male and female patients and levels of severity.

We also investigated the proportion of missing laboratory values over different time intervals to capture the differences in patterns of laboratory missingness. To identify initial differences in missingness during the early days of hospital admission, we plotted on a heatmap the difference in proportion missing during the first 3 days of admission between severe and non-severe patients.

To identify changes in trends of missingness over time, we first modeled the rates of change of proportion missing over time for severe and non-severe patients separately and then plotted on a heatmap the difference in the rates of change. We obtained rates of change by fitting linear models across days since admission and obtaining the beta coefficients. We examined three different time intervals: 0-10 days, 0-30 days, and 0-60 days to capture patients with short, medium, and long term hospital stays. For 0-10 days and 0-30 days, linear models were fit across the proportion missing on each hospital day following admission for all patients. For 0-60 days, the models were fit over three-day periods.

### Identifying relationships between variables based on missingness and ordering patterns

We also characterized the relationships between the labs themselves based on their missingness patterns. To this end, we employed Latent Dirichlet Allocation (LDA) topic modeling to identify similar labs based on their ordering and missingness patterns. LDA is a generative statistical model that allows observations to be explained by latent factors or groups that represent why aspects of the data are similar [42]. It is frequently used in natural language processing where words are observations in a collection of documents, and each document is a mixture of a small number of topics. In LDA, the presence of a given word can be attributed to one of the document’s topics, and topics are determined by words that frequently co-occur. The topic is interpreted to represent a theme in a document. In our setting, we treated the name of each laboratory test as a “word”, each patient admission (up to 30 days) as a document, and our topics are groups of laboratory tests. Our goal was to identify the topics, or “themes”, of labs that frequently occur in our COVID-19 patients across numerous sites.

The input to our LDA topic modeling algorithm is the count of the number of days a laboratory test was ordered up to the first 30 days of a patient’s admission. We first identified the optimal number of topics for our input data; and then LDA learns *β*, or the probabilities for each topic that a given word belongs to a topic; and *θ*, for each patient the probability that the patient contains each topic. For this work, we focus on *β*.

We used four metrics to determine the optimal number of topics to learn from the data; in order to accomplish this, we evaluated a range of two to eight topics. We assessed which of these maximized:

- the held-out likelihood, which provides a measure of how predictive the model is on unseen documents;
- semantic coherence, which captures the tendency of a topic’s high probability words to co-occur in the same document;
- the lower bound on the marginal likelihood;

and which minimized the residuals.

Because every laboratory test has a non-zero probability of belonging to a given topic, we determine a cut-off based on the sum of the cumulative probabilities in decreasing order. If the difference between 1 and the sum of the cumulative probabilities is <= 0.05, then only the labs that have been summed up to that cutoff are determined to describe the topic or make up the majority of the probability. In essence, the labs that make up a topic should be responsible for about 95% of the probability mass.

For each site, we generate a list of topics and the labs that describe that given topic based on their cumulative sum cut-off. From there, we are interested in groups of laboratory tests that intersect topics frequently across sites. We looked for the largest unique combination of labs that intersect at least nine times across all the topics from all the sites. Nine intersections was chosen empirically because it allowed for four unique groups of labs across the 15 sites. From there, we identified other lab tests that might intersect with a given established unique group at least six times to show the heterogeneity between topics.

Lastly, we were interested in the temporal relationships between the various labs based on their missingness. For each lab, we compiled a list of missing indicators for each patient on each day since admission. We then calculated the Spearman correlation between two laboratory test pairs at a given time point based on the missing indicators. We repeated this out to 30 days after admission. Then, for each laboratory test pair, we fit a linear model across the Spearman correlation values across time points. A positive slope indicates that tests were not initially ordered or missing together and they become more concordant as the hospital admission continues; a negative slope indicates that tests are initially ordered or missing together and lose their concordance over time. Both of these dynamics could reveal biological and clinical mechanisms at work.

## Results

### 1. Patterns of missingness each laboratory test across sites

**Figure 1** shows that the missingness across labs varies widely across sites and lab measures. The larger variability in the number of missing values per patient **Figure 1 (A)** compared to the average proportion missing per patient **Figure 1 (B)** across sites is possibly due to differences in the distribution of the length of stay and sample size at particular sites. Nonetheless, the number of missing values per patient still informs the exact number of missing tests and is useful for identifying sites with more valid values which might be required for some analyses. To account for the length of admission, the proportion missing (**Figure 1 (B)**) is normalized against the total number of admitted days for each patient. Generally, creatinine and leukocytes show the lowest number and proportion missing compared to all other tests. This is closely followed by bilirubin, albumin, ALT, and AST, which have a considerably lower number of average missing values per patient and a lower proportion missing (throughout patient admission) across most of the sites. Lymphocytes and neutrophils show a similar level of missingness as the above labs except for a visibly higher measure of missingness in Site 5.

**Figure 1:**
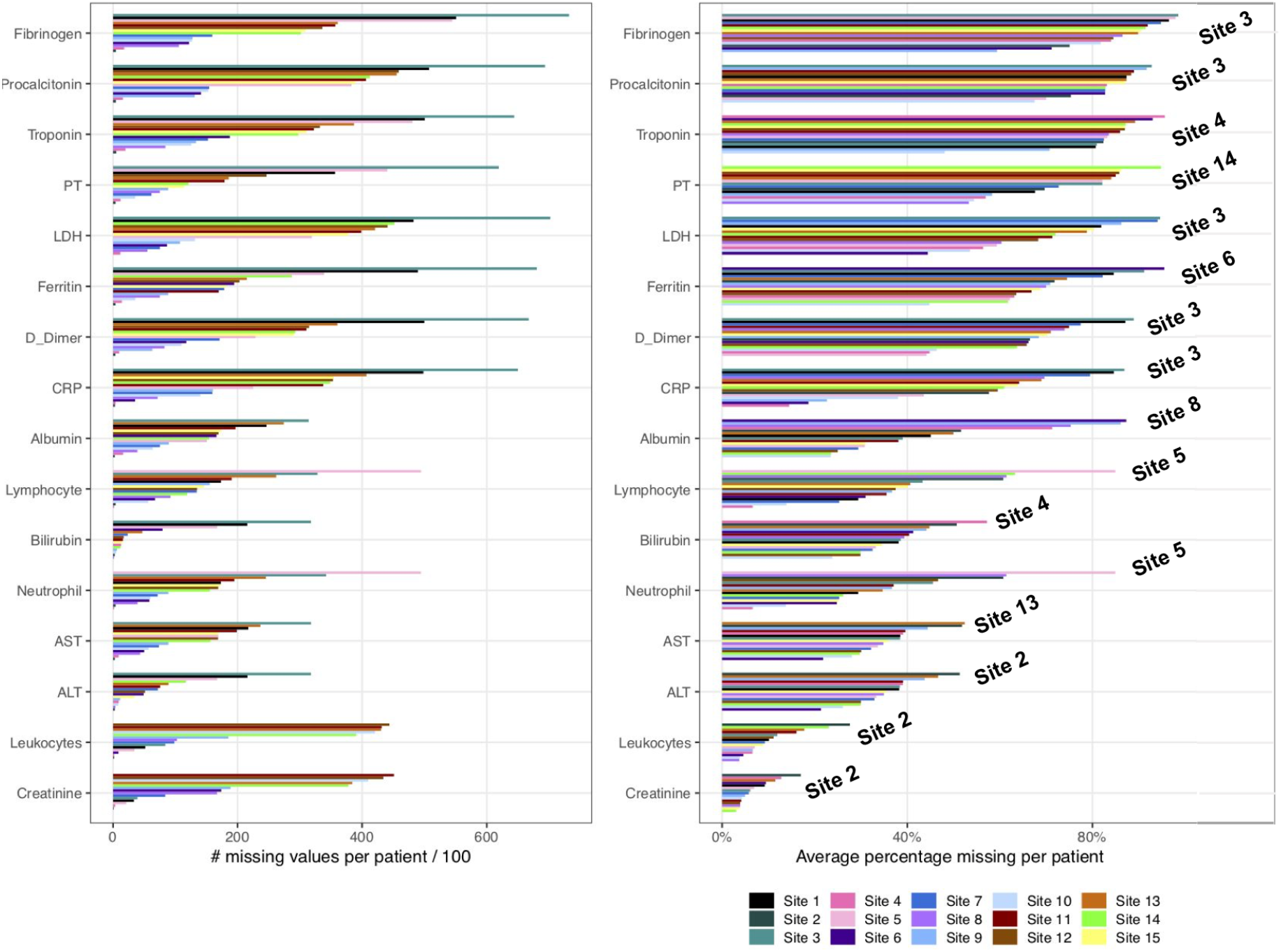
Summary of missingness for lab measures across all sites. (A) Number of missing labs is calculated for each patient before it is averaged over the total number of patients. (B) Proportion missing is calculated by taking into account the total number of days with no measurements of labs divided by the total number of admitted days for each patient. The proportion missing is averaged across all patients. The sites with the highest percentage missing are labeled on the right within the plot.

**Figure 2(A)** shows the difference in proportion missing between males and females for each lab. Generally, across all labs, the difference in missingness by sex is varied in both directions across all sites. We observe the largest difference in missingness between males and females at Site 2, with some female patients having much more missingness for D-dimer, PT, and leukocytes. We also observe some slight deviations at some of the sites with more missingness among females in neutrophils at Site 12 and more missingness among males at Site 13 in AST and bilirubin. It is important to note that Site 2 has the smallest sample size (n=162), and Sites 11-15 have very few female patients (ranging from 5.2-6.4 percent). Thus deviations are likely a result of variability in the data. Beyond these findings, we observe that most sites are well-balanced across the different sexes with regard to missingness in the data.

**Figure 2:**
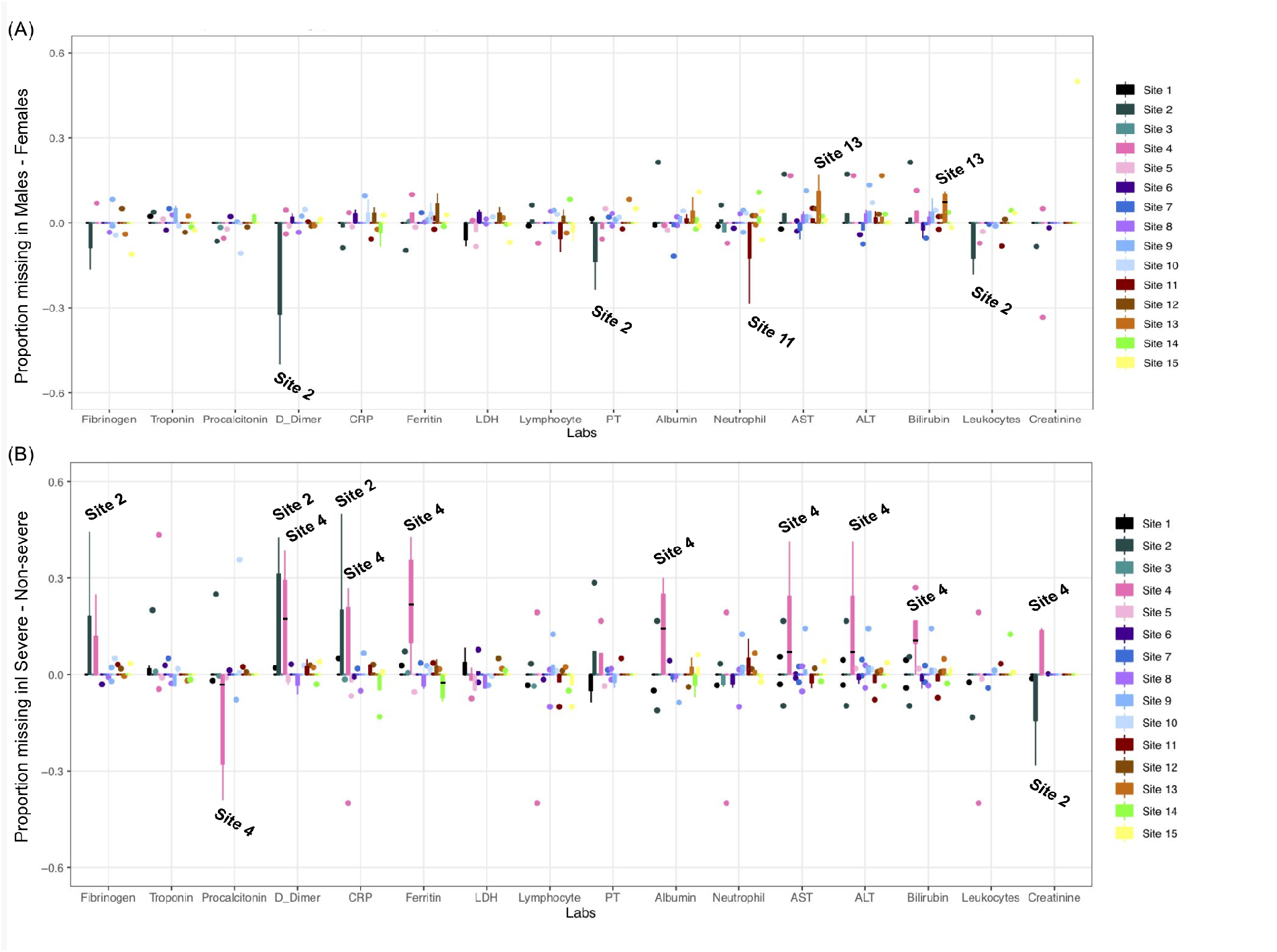
Difference in the quantiles of proportion missing across labs between (A) males and females as well as between (B) severe and non-severe patients. Sites with the largest deviation in the proportion missing are labeled in the plots. For (A), sites with more missingness in the positive direction have more missingness in male populations. For (B), sites with more missingness in the positive direction have more missingness in severe populations.

### 2. Patterns of missingness of laboratory tests by patient severity

Next, we quantified the difference in proportion missing between severe and non-severe patients across the entire cohort in **Figure 2(B)**. We hypothesized for this experiment that there would be a higher proportion missing in non-severe patients. We observe in **Figure 2(B)** that most deviations occur at Site 2 and Site 4. We note more missing data in severe patients at Site 2 for fibrinogen, D-dimer, and CRP, and more missing procalcitonin results in non-severe patients. These attributes could be explained by the fact that Site 2 has a smaller sample size and is a pediatric hospital, hence the current severity definition might not be suited for its patients. At Site 4, we observe more missing data in severe patients for D-dimer, CRP, ferritin, albumin, AST, ALT, bilirubin, and creatinine while we observe more missing procalcitonin data in non-severe patients. Site 4 does not have an intensive care unit (ICU) which could explain the higher proportion of missingness in severe patients. Other than these two sites, the remaining sites seem to be well-balanced across levels of severity with regard to missingness in the data over the whole admission period.

**Figure 3** shows the difference in proportion missing between severe and non-severe patients for the initial three days of admission. For the majority of the sites, we see more missingness in the non-severe group as opposed to the severe group. We see most deviations from this trend in Site 4, with more missingness in the severe group; and a variation of trends at Sites 2 and 9.

**Figure 3:**
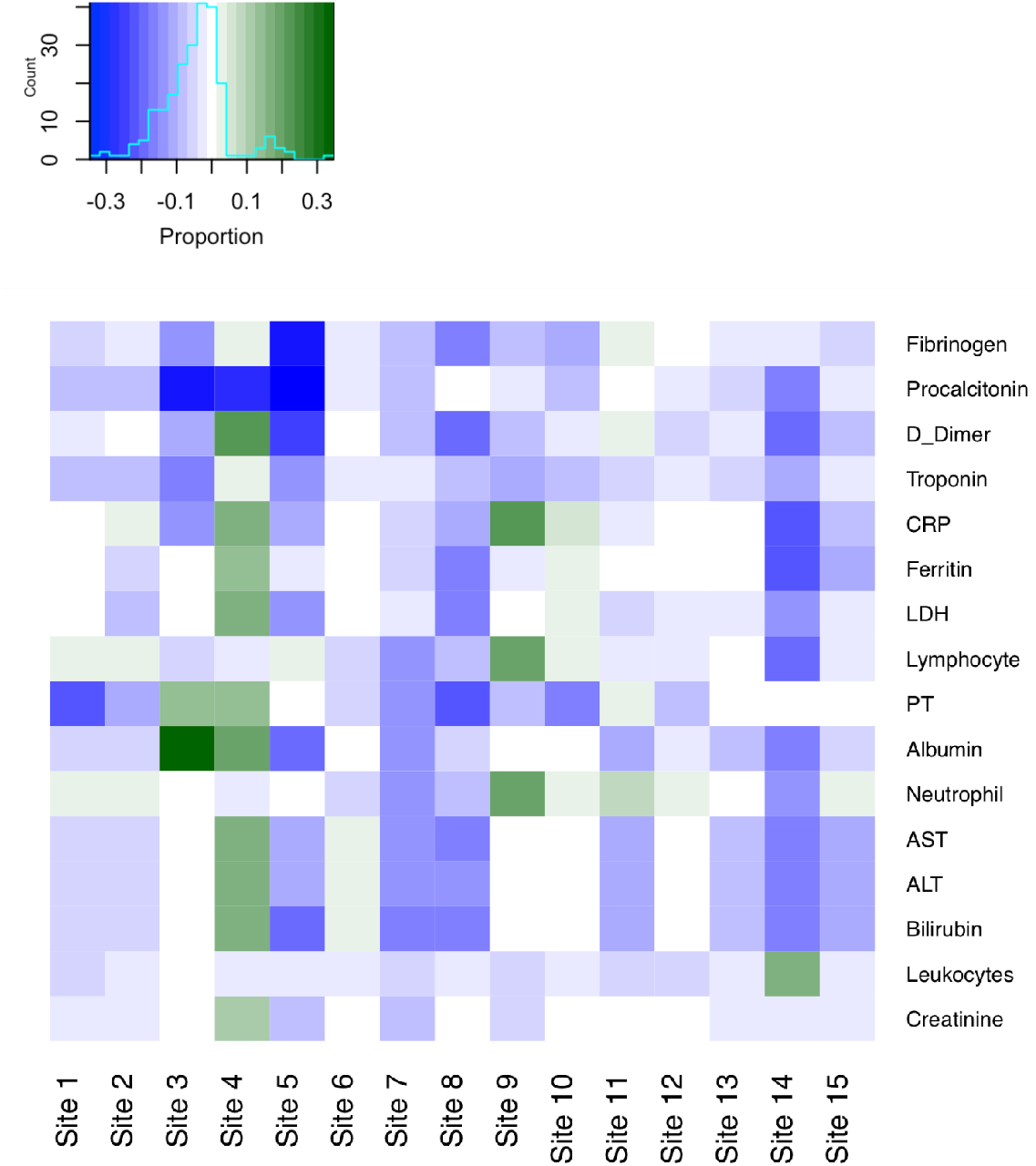
Proportion missing for initial three days of admission. Blue cells represent labs with a higher proportion missing in non-severe patients compared to severe patients. Green cells represent labs with a higher proportion missing in severe patients compared to non-severe patients.The inset indicates the legend for the heatmap and the light blue line represents the counts for each of the difference in proportion missing between severe and non-severe patients.

To understand the differences in proportion missing across admission days, we first modeled the proportion missing across admission days and retrieved the rates of change (beta values) for both severe (**β**_severe_) and non-severe patients (**β**_non-severe_) separately. Then, we took the difference in the rates of change in proportion missing for severe and non-severe patients to draw up the heatmaps across sites and laboratory tests (**Figure 4**). We considered temporal trends in three different time frames: ten days and 30 days for the short-stay patients and 60 days for the long-stay patients. Investigating the trends of missingness across different time frames allows for the visualization of trends that are specific to the time scales.

**Figure 4:**
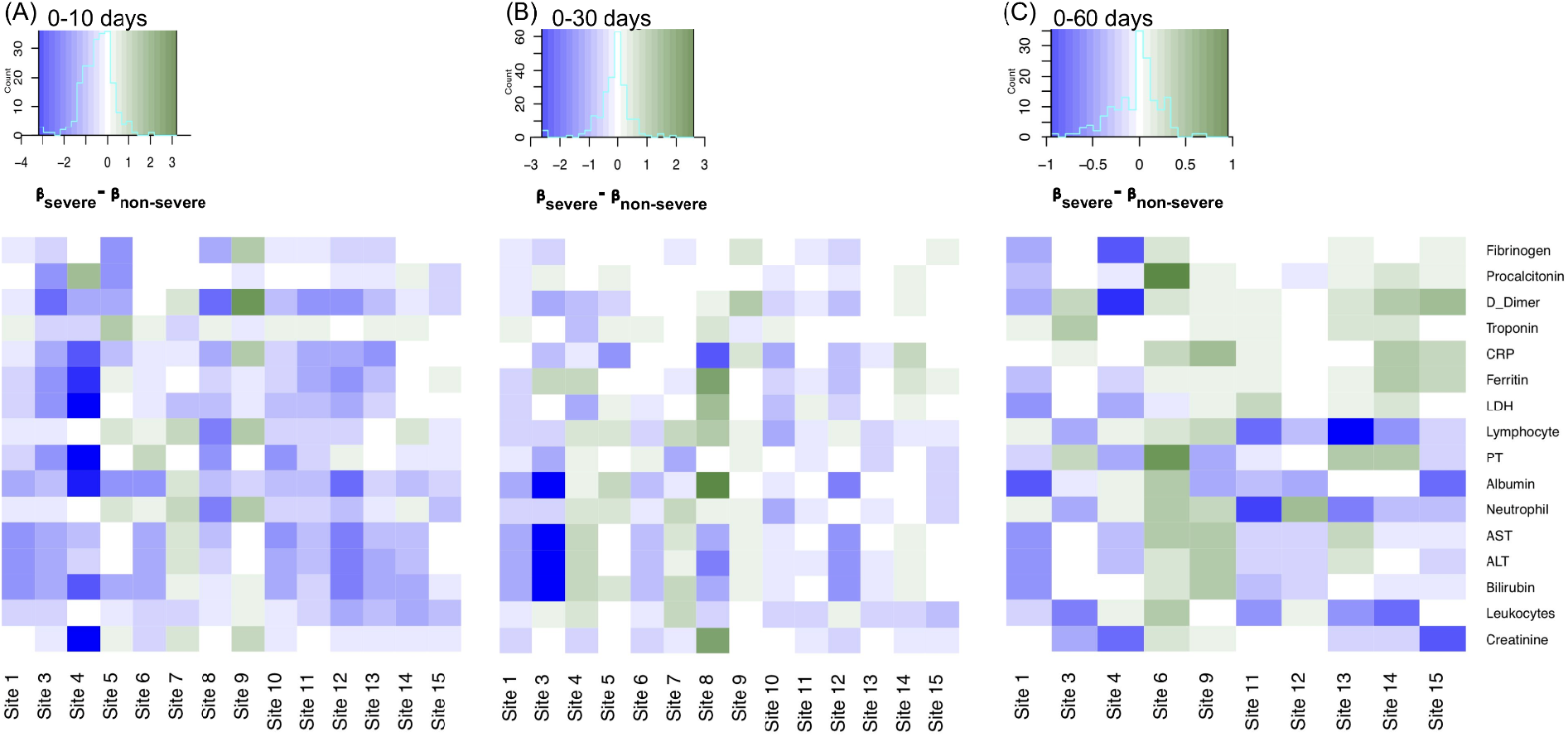
Difference in rates of change in proportion missing in severe and non-severe patients across various time scales. The heatmaps cover temporal changes in rate of change in proportion missing between severe and non-severe patients over the span of (A) 0-10 days (B) 0-30 days and (C) 0-60 days for long-stay patients. Blue cells represent labs with a greater increase in proportion missing in non-severe patients compared to severe patients. Green cells represent labs with a greater increase in proportion missing in severe patients compared to non-severe patients.

From 0-10 days, we observe that while most sites show a higher rate of change in proportion missing in non-severe patients (blue cells), Sites 6, 7, and 9 had multiple labs that show the opposite trend with a higher rate of change in severe patients (shown in green) (**Figure 4(A)**). Site 4 also stood out with a much more pronounced difference, with a much higher rate of change in proportion missing in non-severe patients compared to severe patients (**Figure 4(A)**).

From 0-30 days, we observe a similar trend with a higher rate of change in proportion missing in non-severe patients across the same sites with the addition of Site 4. Also, Site 8 showed a varied, but pronounced difference in the rate of proportion missing across some labs (Ferritin, LDH, Lymphocytes, Albumin, Creatinine) whilst showing a higher rate of change in proportion missing in non-severe patients whilst other labs (CRP, AST, ALT, Bilirubin, Leukocytes) showed a higher rate of change in proportion missing in severe patients (**Figure 4(B)**).

For the long-stay analysis, we included only sites that provided data for patients with hospital stays of at least 60 days. For this, we required sites to have at least three patients with data through 60 hospital days. This led to a shortlist of 10 sites having sufficient data (at least three patients) for this analysis. **Figure 4(C)** that most sites and labs show a greater rate of change in severe patients compared to non-severe patients (green cells). This is with the exception of Sites 1 and 4 with labs that are showing a greater rate of change in proportion in non-severe patients (**Figure 4(C)**). The following temporal line plots display the actual proportions of missingness at various time points for severe and non-severe patients for several labs and can help explain some of these findings.

For example, Troponin also shows a higher rate of change in severe patients across many sites across all time scales. One trend might be that a certain lab has a consistently higher rate of change in severe patients as opposed to non-severe patients (aka mostly green throughout the heatmap in the different time intervals) (**Figure 4**). We see that this is the case for Troponin, as shown in **Figure 5**. Troponin initially has more missingness than all other labs. Our range for possible missingness proportions is limited to [0,1] and initially we see more missingness in the non-severe group as opposed to the severe group. Thus, many sites hit a maximum proportion of missingness much faster in the non-severe group, making the rate of change smaller than it would be for severe patients. We also see more variability in the non-severe group out to later days in the hospital admission.

**Figure 5.**
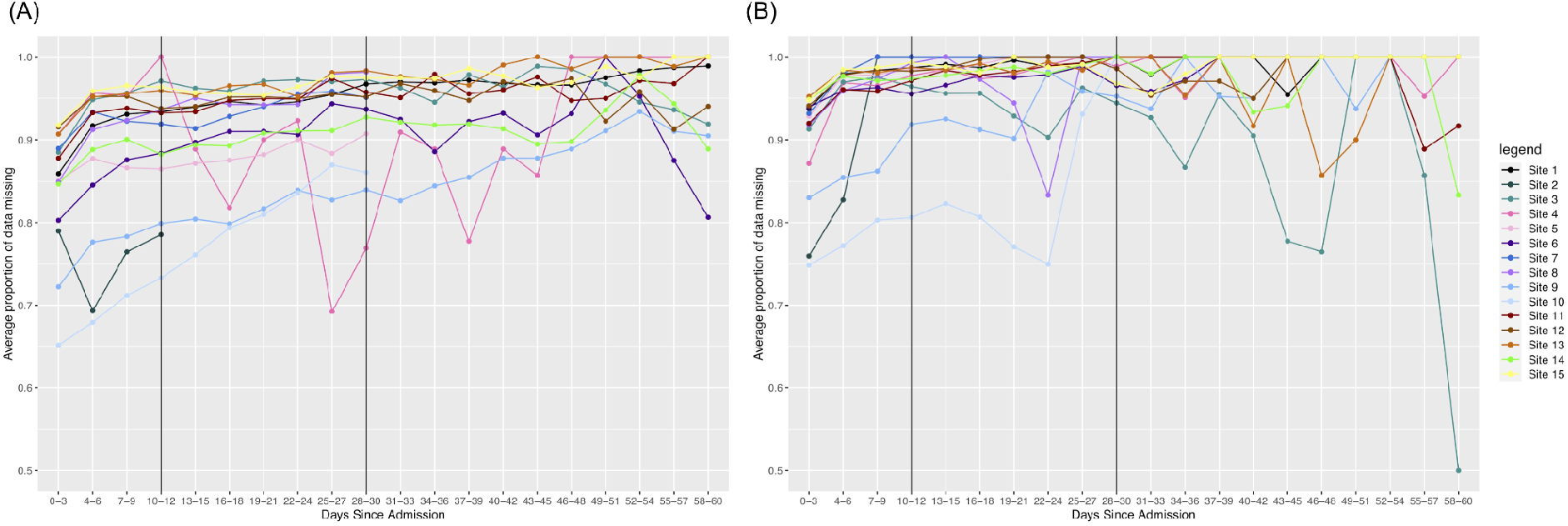
Changes in missing Troponin results over time. (A) severe patients and (B) non-severe patients. Since the range of proportions is limited to [0,1], and non-severe patients initially have more missingness than severe patients, we see that the rate of change across all time is higher for severe patients.

Another trend is that a test may initially have a higher rate of change in non-severe patients early on, but the rate of change is higher in severe patients out to 60 days. We see that this is the case for Ferritin, as shown in **Figure 6**. Initially, we see a stronger increase in missingness in the first 10 days for non-severe patients and this remains to be the case for many sites in 0-30 days. However, again, patients in the non-severe group reach the maximum level of missingness faster than patients in the severe group. Thus, because the severe patients take more time to reach the maximum level of missingness, many sites will have an overall greater rate of change in the severe group.

**Figure 6.**
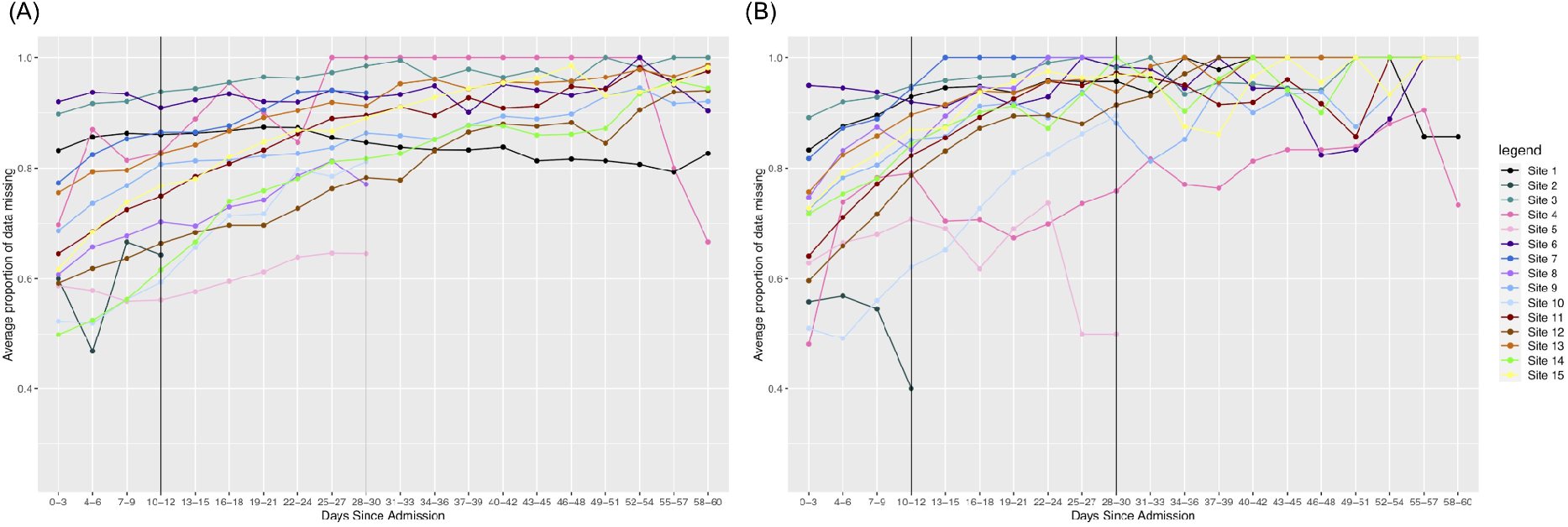
Changes in missing Ferritin results over time. (A) severe patients and (B) non-severe patients. Since the range of proportions is limited to [0,1], and non-severe patients initially have more missingness than severe patients, we see that the rate of change across all time is higher for severe patients.

Lastly, some labs might have a consistently larger rate of change across time intervals for non-severe patients (or mostly blue in the heatmap for the different intervals). We see in **Figure 7** that this is the case for Leukocytes, where this lab has a consistently lower amount of missingness compared to the other labs. Because the upper bound for missingness is further away from the initial amounts of missingness, we see that in non-severe patients it increases more quickly and is more pronounced overall.

**Figure 7.**
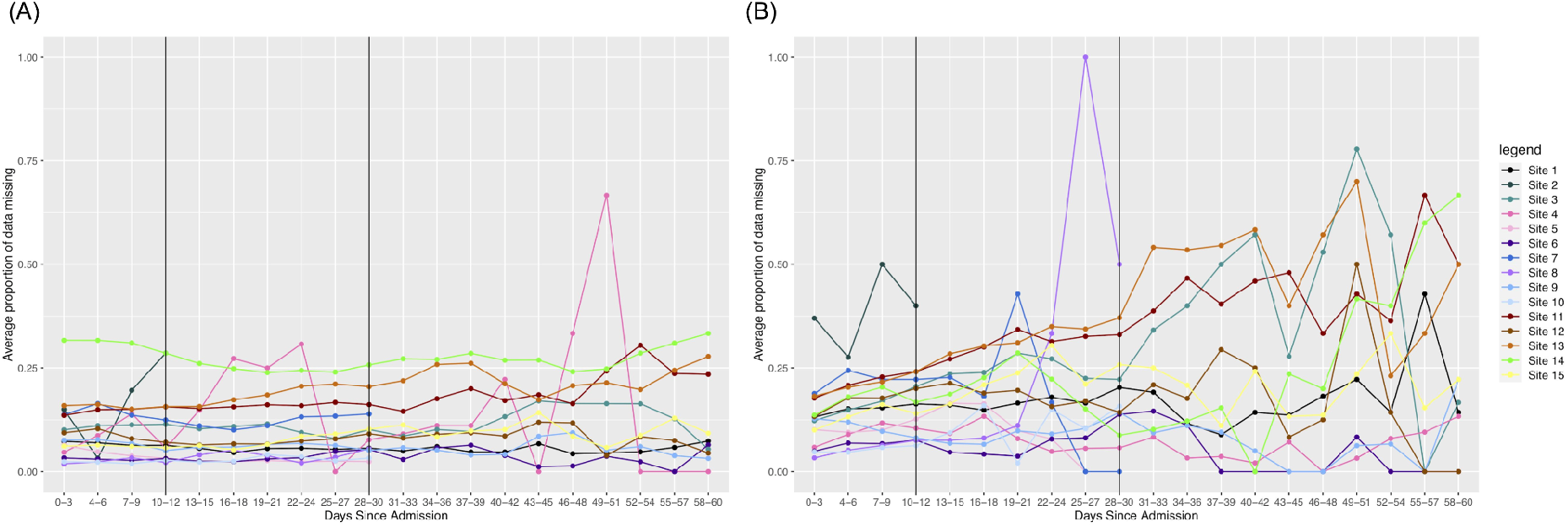
Changes in missing Leukocytes over time. (A) severe patients and (B) non-severe patients. Since the range of proportions is limited to [0,1], and non-severe patients initially have more missingness than severe patients, we see that the rate of change across all time is higher for non-severe patients.

Based on the trends we observe in the heatmaps and some of the temporal line plots, we conclude that overall, there is more missingness in non-severe patients as opposed to severe patients over time across many of the labs in our dataset. This is what we might expect to see because clinicians might not test non-severe patients as heavily over time as severe patients who could be having more issues as their stay continues.

### 3. Patterns of missingness shared between pairs of laboratory tests

In identifying relationships in missingness across labs, we investigated laboratory test pairs that show a change in correlation across admission days (**Figure 8**). We then shortlisted test pairs with either a significant positive or negative change in correlation over admission days (**Figure 8**). Only test pairs that show reproducible correlations across sites are shortlisted. To obtain the rate of change, we fit linear models across the Spearman correlation values over time (**Figure 8 (A) & (B)**). Sites were included for a pair of the slope = |0.3| and standard error <= 0.25.

**Figure 8.**
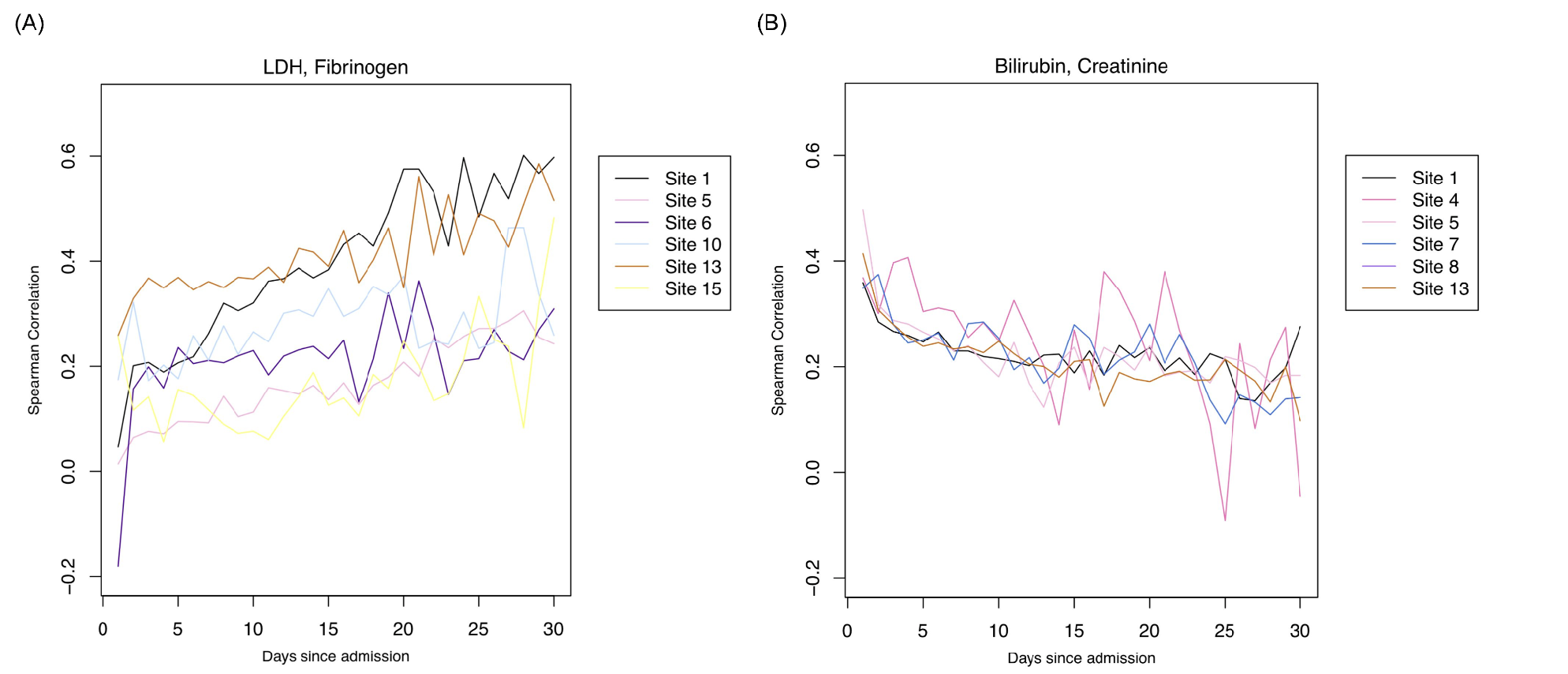
Correlations of pairs of laboratory tests. Pairs of labs with (A) positive or (B) negative correlations through admission. Labs with position correlations are more strongly correlated in their missingness during later parts of admission; Labs with a negative association are more strongly correlated in their missingness during the earlier parts of admission.

In **Table 2**, we observe several lab pairs that become positively associated during the later parts of admission. Notable lab pairs indicate suspicion of infection, coagulopathy including cardiac involvement, liver involvement, severe COVID-19 outcomes as well as rule out of differential diagnoses such as bacterial pneumonia.

**Table 2.** Lab pairs that are more strongly associated during later parts of admission and their implications.

| Pair of Labs | # of Sites | Median Slope (Min, Max) | Implication |
| --- | --- | --- | --- |
| LDH, Fibrinogen | 6 | 0.591 (0.439, 1.608) | Suspicion of infection |
| Fibrinogen, D-dimer | 6 | 0.629 (0.4, 1.843) | Suspicion of coagulopathy |
| Fibrinogen, Procalcitonin | 7 | 0.489 (0.341, 1.6) | Suspicion of coagulopathy with cardiac involvement |
| LDH, Albumin | 6 | 0.5725 (0.33, 0.918) | Suspicion of severe COVID-related outcome |
| LDH, Bilirubin | 6 | 0.6685 (0.347, 0.882) | Suspicion of liver involvement |
| LDH, ALT | 6 | 0.572 (0.348, 0.698) | Suspicion of liver involvement |
| LDH, AST | 6 | 0.541 (0.342, 0.688) | Suspicion of liver involvement |
| Procalcitonin, Ferritin | 7 | 0.698 (0.364, 1.787) | Differential diagnosis- COVID-pneumonia vs. bacterial pneumonia |
| Procalcitonin, D-dimer | 6 | 0.7325 (0.657, 1.7) | Suspicion of severe COVID-related outcome |

In **Table 3**, we observe several lab pairs that become strongly associated during early admission. Notable lab pairs indicate suspicion of infection with or without kidney or liver involvement.

**Table 3.** Lab pairs that are more strongly associated during earlier parts of admission and their implications.

| Pair of Labs | # of Sites | Median Slope (Min, Max) | Implication |
| --- | --- | --- | --- |
| Bilirubin, Creatinine | 6 | -0.5065 (-0.809, -0.311) | Suspicion of kidney involvement |
| ALT, Creatinine | 6 | -0.499 (-0.74, -0.317) | Suspicion of severe COVID-related outcome due to liver and kidney involvement |
| Neutrophil, Creatinine | 6 | -0.44 (-1, -0.31) | Suspicion of infection with kidney involvement |
| Lymphocyte, Creatinine | 6 | -0.5535 (-0.994, -0.307) | Suspicion of infection with kidney involvement |

### 4. Patterns of missingness shared between groups of laboratory tests

To identify groups of labs that share patterns in missingness, topic modeling was done to arrive at a common set of labs across sites (**Figure 9(A)**). From 15 sites, we end up with 91 topics derived from LDA. We include unique groups of labs with at least nine intersections across topics in an pSet plot. Based on these criteria, we obtained four unique groups that are reproducible across sites presented in **Figure 9(A)**. The black points represent the labs that are consistent across all topics found across sites while the pink and red ones represent labs that are only found in some but not all of the topics across sites (**Figure 9(A)**). We find that overall, the groups of intersections represent groups of labs that measure for similar issues in a COVID-19 setting, and thus are related to one another. Group 1 consists of labs that are commonly ordered together on a daily basis, measuring for renal issues and infection. Group 2 are likely tests ordered individually rather than as a group. They all are labs that have at times been thought to help prognosticate potential severity of COVID. Group 3 represents tests that are ordered together to assess liver function, and Group 4 represents coagulation studies that may or may not be ordered as a group.

**Figure 9:**
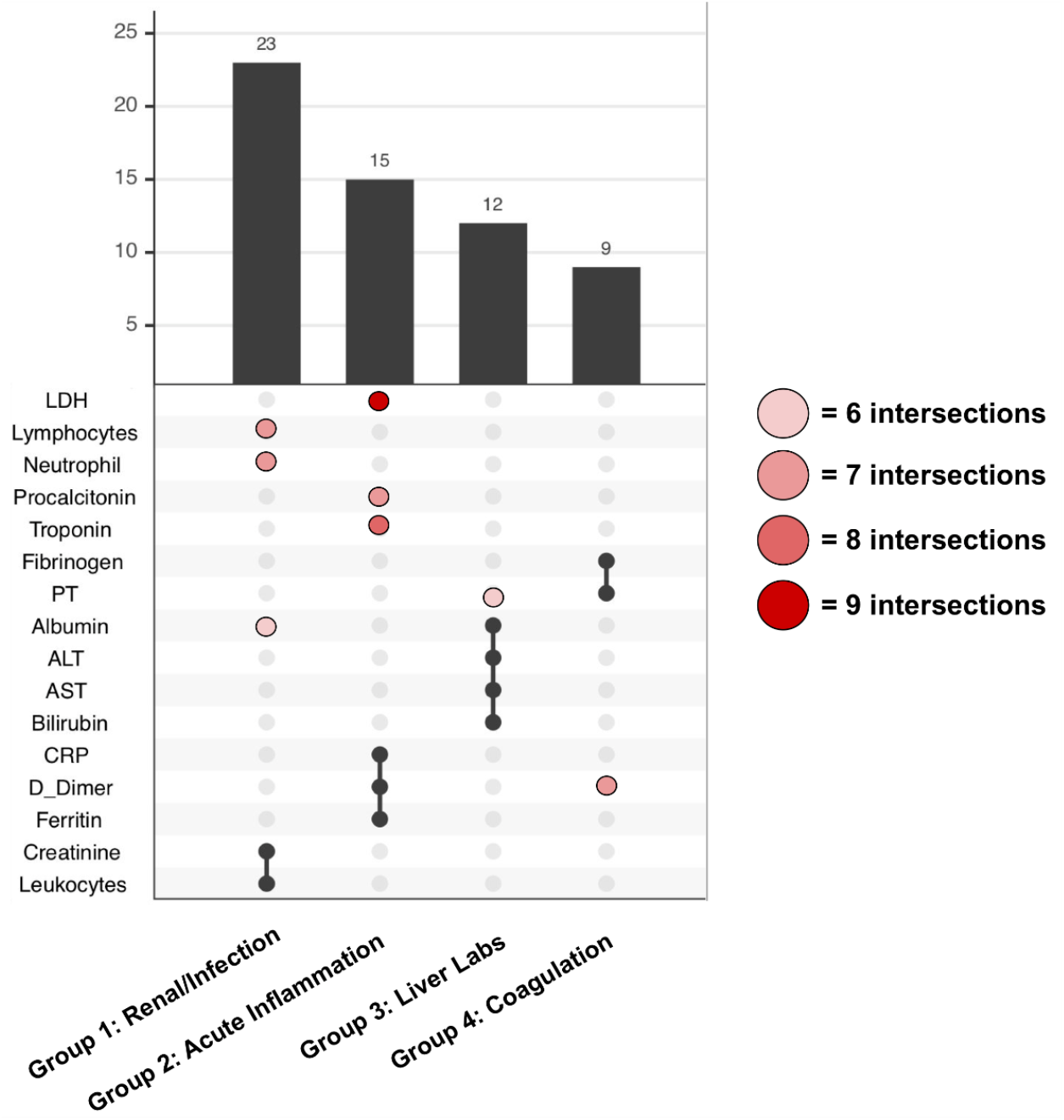
Grouped labs with similar patterns of missingness. Prevalence of the top four groups of labs from topic modeling analyses across sites. The red/pink points show slight deviations in group membership of labs across different sites.

## Discussion

Through characterizing and exploring laboratory test missingness patterns across a multi-national COVID-19 study, we identified several insights and opportunities [1].

### 1. Missingness patterns can indicate data quality issues

Initially, we observed some discrepancies in the average number of missing values in tests that are usually ordered together in the same panel, e.g., Neutrophils and Lymphocytes, in **Figure 1**. The sites that had this discrepancy were Sites 1, 6, and Sites 11 through 15. At Site 1, these findings led to a correction of lab mappings to LOINC codes used at some sites and a subsequent correction in the data. We also found some discrepancies in the proportions of hepatic function tests at Site 13 and Site 10, with slightly more AST measurements as opposed to ALT. Mapping issues can be due to miscoding and issues with LOINC code specificity. Deriving knowledge of laboratory ordering patterns from clinical experts can help with developing more robust local data quality improvement checks. Researchers can work locally with their academic medical centers to provide useful solutions to address these inaccuracies and operationalize these data quality processes at scale.

### 2. Missingness patterns can indicate responses to hospital treatment capacity

During the global pandemic, several academic medical centers were not prepared for the large influx of patients with intensive care needs. Many created makeshift ICUs. Furthermore, some facilities may have implemented new triaging procedures. For example, the higher proportion of missing data in the initial three days of admission in Site 4 across almost all the labs could be attributed to the lack of an ICU in Site 4. Because patients with serious conditions might be in the process of having a transfer being arranged, it could explain the higher proportion of missing labs in the severe patients from Site 4.

### 3. Missingness patterns can change over time and at different rates

Although all patient groups exhibit some proportion of non-reported labs, not all patient groups consistently demonstrate the same rate of non-reported labs over time. Patients among the non-severe group reach the maximum level of non-ordered labs faster than patients among the severe group. For short-stay patients, we observe this change for labs like troponin. For ferritin, we observe a higher rate of change in non-severe patients. Intuitively, for non-severe patients, clinicians may become less concerned about a patient’s condition, leading to reduced ordering rates. As a corollary, the absence of test ordering can indicate improvements in the patient’s health.

### 4. Missingness patterns could be predictive of clinical outcomes

Missingness patterns in healthcare data also carries a signal within itself. Lack of a reported lab could possibly indicate how unimportant the lab is in the progression or monitoring of disease and hence be an important predictor for clinical outcomes as well. Because the reporting patterns between labs are cross-correlated (groups of labs with shared reporting (and non-reporting) patterns as shown in topic modeling **Figure 9**), the removal of one test’s missing values from a model could potentially affect model performance as well. Furthermore, using two different data-driven methods -- spearman correlations over time as well as topic models -- we observed common clinical themes among lab pairs and sets including coagulation, infection with renal involvement, and liver involvement.

### 5. Computational approaches for addressing ordering patterns

Missingness patterns could inform which variables would benefit from imputation for future studies. Including them as variables could be problematic because it is not exactly independent of variables. A number of approaches to dealing with informative missingness have been reported. These include using a Monte Carlo Expectation-Maximization simulation series that incorporates within-subject autocorrelation with a latent autoregressive process for longitudinal binary data [43], a Bayesian shrinkage approach to improve estimation of sparse patterns [44], and the use of an informative ordering pattern odds ratio [45–47]. Continued work in temporal pattern estimation in the face of informative missing data will investigate these methods in the context of these COVID consortium data [48].

This study has several limitations.

- *Order sets were not investigated*. The patterns of missingness for laboratory and other tests and procedures were likely influenced by standing order sets. Furthermore, order sets are likely to change over time as knowledge about COVID and therapies for it evolve over time. We plan to correlate patterns of missingness with the content of order sets and how they change over the course of the pandemic.
- *Missingness was not correlated with secular trends of the pandemic*. As the pandemic has evolved, there have been a number of irregular cycles in the epidemic curve, with marked changes in disease incidence associated with the Delta and Omicron variants. These changes are likely associated with corresponding changes in order sets and clinical practice. We will investigate patterns of laboratory test missingness as they may reflect clinical practice patterns which in turn may reflect the undulation in the incidence of COVID over time.
- *Type of patient care unit was not captured*. It is possible that test ordering patterns could be influenced by the type of unit a patient was on at given points in time. For example, order sets may be used less frequently in intensive care units than in medical units, even those dedicated to COVID. Our future work will include an examination of “unit effect” in a temporal context, which will be important as patients move between acute and less-acute care settings in a given hospitalization.
- *Focus on missing test values, as opposed to missing orders*. We assumed that a missing laboratory test result indicated that the test was not ordered. While a reasonable assumption, a more accurate indicator of ordering behavior would be to capture orders in addition to test results. We will investigate the feasibility of obtaining these data from the EHR in future work.
- *Severity definitions might change over time*. When patients stay in the hospital for long periods of time, it is possible that their severity changes. Thus, there are limitations in looking at **Figure 4** due to the fact that patients initially labeled as non-severe that remain in the hospital out to 60 days might not actually be non-severe at that point.

## Conclusion

In this study, we investigated and demonstrated how characterization of missing data patterns in EHRs, particularly lab results, could support various steps in scientific study ranging from data quality to hypothesis generation. Furthermore, missing data patterns will enable consortia to identify which sites are better poised to study particular questions and potentially inform the use of imputation methods for addressing these challenges. Finally, our results may provide insights into some of the biological relationships between labs in EHRs data for COVID-19 patients.

## Data Availability

All data produced in the present study are available upon reasonable request to the authors.

## Supplementary Materials

**Supplementary Table 1.** Description of laboratory tests, results, and their clinical significance [38,41].

| **Laboratory test** | **Trend** | **Clinical significance** | **Commonly Ordered Inpatient Labs** |
| --- | --- | --- | --- |
| Prothrombin Time (PT) | Increased | Bleeding disorders | Yes |
| Aspartate Aminotransferase (AST) | Increased | Impaired hepatic function | Yes |
| Alanine Aminotransferase (ALT) | Increased | Impaired hepatic function | Yes |
| Creatinine | Increased | Impaired renal function | Yes |
| Albumin | Decreased | Impaired hepatic or renal function | Yes |
| Leukocytes | Increased | Infection | Yes |
| Lymphocyte | Increased | Infection | Yes |
| Neutrophil | Increased | Infection | Yes |
| Fibrinogen | Decreased<br>Increased | Bleeding disorders<br>Clotting disorders | No |
| D-dimer | Increased | Clotting disorder | No |
| Troponin | Increased | Cardiac injury | No |
| Bilirubin | Increased | Impaired hepatic function | Yes |
| Lactate Dehydrogenase (LDH) | Increased | Liver, lung, and other tissue damage | No |
| Procalcitonin | Increased | Infection, sepsis | No |
| Ferritin | Increased | Inflammation, infection | No |
| C-Reactive Protein (CRP) | Increased | Inflammation | No |

## References

[1] J.C. Denny, Chapter 13: Mining electronic health records in the genomics era, PLoS Comput. Biol. 8 (2012) e1002823.

[2] R.A. Bush, C.D. Connelly, A. Pérez, H. Barlow, G.J. Chiang, Extracting autism spectrum disorder data from the electronic health record, Appl. Clin. Inform. 8 (2017) 731–741.

[3] M. Apte, M. Neidell, E.Y. Furuya, D. Caplan, S. Glied, E. Larson, Using electronically available inpatient hospital data for research, Clin. Transl. Sci. 4 (2011) 338–345.

[4] M.S. Dittmar, S. Zimmermann, M. Creutzenberg, S. Bele, D. Bitzinger, D. Lunz, B.M. Graf, M. Kieninger, Evaluation of comprehensiveness and reliability of electronic health records concerning resuscitation efforts within academic intensive care units: a retrospective chart analysis, BMC Emerg. Med. 21 (2021) 69.

[5] R. Farmer, R. Mathur, K. Bhaskaran, S.V. Eastwood, N. Chaturvedi, L. Smeeth, Promises and pitfalls of electronic health record analysis, Diabetologia. 61 (2018) 1241– 1248.

[6] K.B. Bayley, T. Belnap, L. Savitz, A.L. Masica, N. Shah, N.S. Fleming, Challenges in using electronic health record data for CER: experience of 4 learning organizations and solutions applied, Med. Care. 51 (2013) S80–6.

[7] L. Samal, P.C. Dykes, J.O. Greenberg, O. Hasan, A.K. Venkatesh, L.A. Volk, D.W. Bates, Care coordination gaps due to lack of interoperability in the United States: a qualitative study and literature review, BMC Health Serv. Res. 16 (2016) 143.

[8] H. Aerts, D. Kalra, C. Sáez, J.M. Ramírez-Anguita, M.-A. Mayer, J.M. Garcia-Gomez, M. Durà-Hernández, G. Thienpont, P. Coorevits, Quality of Hospital Electronic Health Record (EHR) Data Based on the International Consortium for Health Outcomes Measurement (ICHOM) in Heart Failure: Pilot Data Quality Assessment Study, JMIR Med Inform. 9 (2021) e27842.

[9] M.Y. Argalious, J.E. Dalton, T. Sreenivasalu, J. O’Hara, D.I. Sessler, The association of preoperative statin use and acute kidney injury after noncardiac surgery, Anesth. Analg. 117 (2013) 916–923.

[10] C. Chang, Y. Deng, X. Jiang, Q. Long, Multiple imputation for analysis of incomplete data in distributed health data networks, Nat. Commun. 11 (2020) 5467.

[11] S.S. Feldman, G. Davlyatov, A.G. Hall, Toward Understanding the Value of Missing Social Determinants of Health Data in Care Transition Planning, Appl. Clin. Inform. 11 (2020) 556–563.

[12] M.A. Gianfrancesco, N.D. Goldstein, A narrative review on the validity of electronic health record-based research in epidemiology, BMC Med. Res. Methodol. 21 (2021) 234.

[13] B.O. Petrazzini, H. Naya, F. Lopez-Bello, G. Vazquez, L. Spangenberg, Evaluation of different approaches for missing data imputation on features associated to genomic data, BioData Min. 14 (2021) 1–13.

[14] C.J. Sammon, A. Miller, K.R. Mahtani, T.A. Holt, N.J. McHugh, R.A. Luqmani, A.L. Nightingale, Missing laboratory test data in electronic general practice records: analysis of rheumatoid factor recording in the clinical practice research datalink, Pharmacoepidemiol. Drug Saf. 24 (2015) 504–509.

[15] Z. Che, S. Purushotham, K. Cho, D. Sontag, Y. Liu, Recurrent Neural Networks for Multivariate Time Series with Missing Values, Sci. Rep. 8 (2018) 6085.

[16] A.S. Allen, J.S. Collins, P.J. Rathouz, C.L. Selander, G.A. Satten, Bootstrap calibration of TRANSMIT for informative missingness of parental genotype data, BMC Genet. 4 Suppl 1 (2003) S39.

[17] A.S. Allen, P.J. Rathouz, G.A. Satten, Informative missingness in genetic association studies: case-parent designs, Am. J. Hum. Genet. 72 (2003) 671–680.

[18] I. James, E. McKinnon, S. Gaudieri, G. Morahan, Diabetes Genetics Consortium, Missingness in the T1DGC MHC fine-mapping SNP data: association with HLA genotype and potential influence on genetic association studies, Diabetes Obes. Metab. 11 Suppl 1 (2009) 101–107.

[19] M. Kujala, J. Nevalainen, A case study of normalization, missing data and variable selection methods in lipidomics, Stat. Med. 34 (2015) 59–73.

[20] W.-Y. Lin, N. Liu, Reducing bias of allele frequency estimates by modeling SNP genotype data with informative missingness, Front. Genet. 3 (2012) 107.

[21] S.H. Liu, G. Erion, V. Novitsky, V. De Gruttola, Viral Genetic Linkage Analysis in the Presence of Missing Data, PLoS One. 10 (2015) e0135469.

[22] N.M. Butera, D. Zeng, A. Green Howard, P. Gordon-Larsen, J. Cai, A doubly robust method to handle missing multilevel outcome data with application to the China Health and Nutrition Survey, Stat. Med. 41 (2022) 769–785.

[23] M.C. Wu, D.A. Follmann, Use of summary measures to adjust for informative missingness in repeated measures data with random effects, Biometrics. 55 (1999) 75– 84.

[24] A. Chaimani, D. Mavridis, J.P.T. Higgins, G. Salanti, I.R. White, Allowing for informative missingness in aggregate data meta-analysis with continuous or binary outcomes: Extensions to metamiss, Stata J. 18 (2018) 716–740.

[25] R.G. Harris, M. Batterham, E.P. Neale, I. Ferreira, Impact of missing outcome data in meta-analyses of lifestyle interventions during pregnancy to reduce postpartum weight retention: An overview of systematic reviews with meta-analyses and additional sensitivity analyses, Obes. Rev. 22 (2021) e13318.

[26] L.A. Kahale, A.M. Khamis, B. Diab, Y. Chang, L.C. Lopes, A. Agarwal, L. Li, R.A. Mustafa, S. Koujanian, R. Waziry, J.W. Busse, A. Dakik, H.J. Schünemann, L. Hooft, R.J. Scholten, G.H. Guyatt, E.A. Akl, Potential impact of missing outcome data on treatment effects in systematic reviews: imputation study, BMJ. (2020) m2898. https://doi.org/10.1136/bmj.m2898.

[27] D. Mavridis, G. Salanti, T.A. Furukawa, A. Cipriani, A. Chaimani, I.R. White, Allowing for uncertainty due to missing and LOCF imputed outcomes in meta-analysis, Stat. Med. 38 (2019) 720–737.

[28] D. Mavridis, I.R. White, J.P.T. Higgins, A. Cipriani, G. Salanti, Allowing for uncertainty due to missing continuous outcome data in pairwise and network meta-analysis, Stat. Med. 34 (2015) 721–741.

[29] I.R. White, J.P.T. Higgins, A.M. Wood, Allowing for uncertainty due to missing data in meta-analysis--part 1: two-stage methods, Stat. Med. 27 (2008) 711–727.

[30] R.H. Lyles, A.S. Allen, W. Dana Flanders, L.L. Kupper, D.L. Christensen, Inference for case-control studies when exposure status is both informatively missing and misclassified, Statistics in Medicine. 25 (2006) 4065–4080. https://doi.org/10.1002/sim.2500.

[31] R.H.H. Groenwold, Informative missingness in electronic health record systems: the curse of knowing, Diagn Progn Res. 4 (2020) 8.

[32] G.A. Brat, G.M. Weber, N. Gehlenborg, P. Avillach, N.P. Palmer, L. Chiovato, J. Cimino, L.R. Waitman, G.S. Omenn, A. Malovini, J.H. Moore, B.K. Beaulieu-Jones, V. Tibollo, S.N. Murphy, S.L. Yi, M.S. Keller, R. Bellazzi, D.A. Hanauer, A. Serret-Larmande, A. Gutierrez-Sacristan, J.J. Holmes, D.S. Bell, K.D. Mandl, R.W. Follett, J.G. Klann, D.A. Murad, L. Scudeller, M. Bucalo, K. Kirchoff, J. Craig, J. Obeid, V. Jouhet, R. Griffier, S. Cossin, B. Moal, L.P. Patel, A. Bellasi, H.U. Prokosch, D. Kraska, P. Sliz, A.L.M. Tan, K.Y. Ngiam, A. Zambelli, D.L. Mowery, E. Schiver, B. Devkota, R.L. Bradford, M. Daniar, C. Daniel, V. Benoit, R. Bey, N. Paris, P. Serre, N. Orlova, J. Dubiel, M. Hilka, A.S. Jannot, S. Breant, J. Leblanc, N. Griffon, A. Burgun, M. Bernaux, A. Sandrin, E. Salamanca, S. Cormont, T. Ganslandt, T. Gradinger, J. Champ, M. Boeker, P. Martel, L. Esteve, A. Gramfort, O. Grisel, D. Leprovost, T. Moreau, G. Varoquaux, J.-J. Vie, D. Wassermann, A. Mensch, C. Caucheteux, C. Haverkamp, G. Lemaitre, S. Bosari, I.D. Krantz, A. South, T. Cai, I.S. Kohane, International electronic health record-derived COVID-19 clinical course profiles: the 4CE consortium, NPJ Digit Med. 3 (2020) 109.

[33] J.G. Klann, G.M. Weber, H. Estiri, B. Moal, P. Avillach, C. Hong, V. Castro, T. Maulhardt, A.L.M. Tan, A. Geva, B.K. Beaulieu-Jones, A. Malovini, A.M. South, S. Visweswaran, G.S. Omenn, K.Y. Ngiam, K.D. Mandl, M. Boeker, K.L. Olson, D.L. Mowery, M. Morris, R.W. Follett, D.A. Hanauer, R. Bellazzi, J.H. Moore, N.-H.W. Loh, D.S. Bell, K.B. Wagholikar, L. Chiovato, V. Tibollo, S. Rieg, A.L.L.J. Li, V. Jouhet, E. Schriver, M.J. Samayamuthu, Z. Xia, M. Hutch, Y. Luo, Consortium for Clinical Characterization of COVID-19 by EHR (4CE) (CONSORTIA AUTHOR), I.S. Kohane, G.A. Brat, S.N. Murphy, Validation of an Internationally Derived Patient Severity Phenotype to Support COVID-19 Analytics from Electronic Health Record Data, J. Am. Med. Inform. Assoc. (2021). https://doi.org/10.1093/jamia/ocab018.

[34] G.M. Weber, C. Hong, N.P. Palmer, P. Avillach, S.N. Murphy, A. Gutiérrez-Sacristán, Z. Xia, A. Serret-Larmande, A. Neuraz, G.S. Omenn, S. Visweswaran, J.G. Klann, A.M. South, N.H.W. Loh, M. Cannataro, B.K. Beaulieu-Jones, R. Bellazzi, G. Agapito, M. Alessiani, B.J. Aronow, D.S. Bell, A. Bellasi, V. Benoit, M. Beraghi, M. Boeker, J. Booth, S. Bosari, F.T. Bourgeois, N.W. Brown, M. Bucalo, L. Chiovato, L. Chiudinelli, A. Dagliati, B. Devkota, S.L. DuVall, R.W. Follett, T. Ganslandt, N. García Barrio, T. Gradinger, R. Griffier, D.A. Hanauer, J.H. Holmes, P. Horki, K.M. Huling, R.W. Issitt, V. Jouhet, M.S. Keller, D. Kraska, M. Liu, Y. Luo, K.E. Lynch, A. Malovini, K.D. Mandl, C. Mao, A. Maram, M.E. Matheny, T. Maulhardt, M. Mazzitelli, M. Milano, J.H. Moore, J.S. Morris, M. Morris, D.L. Mowery, T.P. Naughton, K.Y. Ngiam, J.B. Norman, L.P. Patel, M. Pedrera Jimenez, R.B. Ramoni, E.R. Schriver, L. Scudeller, N.J. Sebire, P. Serrano Balazote, A. Spiridou, A.L. Tan, B.W.L. Tan, V. Tibollo, C. Torti, E.M. Trecarichi, M. Vitacca, A. Zambelli, C. Zucco, I.S. Kohane, T. Cai, G.A. Brat, International Comparisons of Harmonized Laboratory Value Trajectories to Predict Severe COVID-19: Leveraging the 4CE Collaborative Across 342 Hospitals and 6 Countries: A Retrospective Cohort Study, medRxiv. (2021). https://doi.org/10.1101/2020.12.16.20247684.

[35] T.T. Le, A. Gutiérrez-Sacristán, J. Son, C. Hong, A.M. South, B.K. Beaulieu-Jones, N.H.W. Loh, Y. Luo, M. Morris, K.Y. Ngiam, L.P. Patel, M.J. Samayamuthu, E. Schriver, A.L.M. Tan, J. Moore, T. Cai, G.S. Omenn, P. Avillach, I.S. Kohane, Consortium for Clinical Characterization of COVID-19 by EHR (4CE), S. Visweswaran, D.L. Mowery, Z. Xia, Multinational characterization of neurological phenotypes in patients hospitalized with COVID-19, Sci. Rep. 11 (2021) 20238.

[36] F.T. Bourgeois, A. Gutiérrez-Sacristán, M.S. Keller, M. Liu, C. Hong, C.-L. Bonzel, A.L.M. Tan, B.J. Aronow, M. Boeker, J. Booth, J. Cruz Rojo, B. Devkota, N. García Barrio, N. Gehlenborg, A. Geva, D.A. Hanauer, M.R. Hutch, R.W. Issitt, J.G. Klann, Y. Luo, K.D. Mandl, C. Mao, B. Moal, K.L. Moshal, S.N. Murphy, A. Neuraz, K.Y. Ngiam, G.S. Omenn, L.P. Patel, M.P. Jiménez, N.J. Sebire, P.S. Balazote, A. Serret-Larmande, A.M. South, A. Spiridou, D.M. Taylor, P. Tippmann, S. Visweswaran, G.M. Weber, I.S. Kohane, T. Cai, P. Avillach, Consortium for Clinical Characterization of COVID-19 by EHR (4CE), International Analysis of Electronic Health Records of Children and Youth Hospitalized With COVID-19 Infection in 6 Countries, JAMA Netw Open. 4 (2021) e2112596.

[37] H. Estiri, Z.H. Strasser, G.A. Brat, Y.R. Semenov, Consortium for Characterization of COVID-19 by EHR (4CE), C.J. Patel, S.N. Murphy, Evolving phenotypes of non-hospitalized patients that indicate long COVID, BMC Med. 19 (2021) 249.

[38] J.W. Rudolf, A.S. Dighe, C.M. Coley, I.K. Kamis, B.M. Wertheim, D.E. Wright, K.B. Lewandrowski, J.M. Baron, Analysis of Daily Laboratory Orders at a Large Urban Academic Center: A Multifaceted Approach to Changing Test Ordering Patterns, Am. J. Clin. Pathol. 148 (2017) 128.

[39] Website, (n.d.). https://shmpublications.onlinelibrary.wiley.com/doi/full/10.1002/jhm.2354?casa_token=Tqb8-Vv7sbgAAAAA%3AT5YCnn2VadQTjLM83pk6_zI2fmn9nzKPCGzzf_KE8SRDoSa1ZyCioazf0eCn6nxV3fuV2bul6FD0DpE.

[40] Journal of Hospital Medicine, (n.d.). https://doi.org/10.1002/(ISSN)1553-5606.

[41] A.H. Corson, V.S. Fan, T. White, S.D. Sullivan, K. Asakura, M. Myint, C.R. Dale, A multifaceted hospitalist quality improvement intervention: Decreased frequency of common labs, J. Hosp. Med. 10 (2015) 390–395.

[42] D.M. Blei, A.Y. Ng, M.I. Jordan, Latent dirichlet allocation, J. Mach. Learn. Res. 3 (2003) 993–1022.

[43] P.S. Albert, D.A. Follmann, S.A. Wang, E.B. Suh, A latent autoregressive model for longitudinal binary data subject to informative missingness, Biometrics. 58 (2002) 631– 642.

[44] J.T. Gaskins, M.J. Daniels, B.H. Marcus, Bayesian methods for nonignorable dropout in joint models in smoking cessation studies, J. Am. Stat. Assoc. 111 (2016) 1454–1465.

[45] J.P.T. Higgins, I.R. White, A.M. Wood, Imputation methods for missing outcome data in meta-analysis of clinical trials, Clin. Trials. 5 (2008) 225–239.

[46] L.M. Spineli, An empirical comparison of Bayesian modelling strategies for missing binary outcome data in network meta-analysis, BMC Med. Res. Methodol. 19 (2019) 86.

[47] L.M. Spineli, C. Kalyvas, K. Pateras, Participants’ outcomes gone missing within a network of interventions: Bayesian modeling strategies, Stat. Med. 38 (2019) 3861– 3879.

[48] G.M. Weber, H.G. Zhang, S. L’Yi, C.-L. Bonzel, C. Hong, P. Avillach, A. Gutiérrez-Sacristán, N.P. Palmer, A.L.M. Tan, X. Wang, W. Yuan, N. Gehlenborg, A. Alloni, D.F. Amendola, A. Bellasi, R. Bellazzi, M. Beraghi, M. Bucalo, L. Chiovato, K. Cho, A. Dagliati, H. Estiri, R.W. Follett, N. García Barrio, D.A. Hanauer, D.W. Henderson, Y.-L. Ho, J.H. Holmes, M.R. Hutch, R. Kavuluru, K. Kirchoff, J.G. Klann, A.K. Krishnamurthy, T.T. Le, M. Liu, N.H.W. Loh, S. Lozano-Zahonero, Y. Luo, S. Maidlow, A. Makoudjou, A. Malovini, M.R. Martins, B. Moal, M. Morris, D.L. Mowery, S.N. Murphy, A. Neuraz, K.Y. Ngiam, M.P. Okoshi, G.S. Omenn, L.P. Patel, M. Pedrera Jiménez, R.A. Prudente, M.J. Samayamuthu, F.J. Sanz Vidorreta, E.R. Schriver, P. Schubert, P. Serrano Balazote, B.W. Tan, S.E. Tanni, V. Tibollo, S. Visweswaran, K.B. Wagholikar, Z. Xia, D. Zöller, Consortium For Clinical Characterization Of COVID-19 By EHR (4CE), I.S. Kohane, T. Cai, A.M. South, G.A. Brat, International Changes in COVID-19 Clinical Trajectories Across 315 Hospitals and 6 Countries: Retrospective Cohort Study, J. Med. Internet Res. 23 (2021) e31400.

